# ROC Analysis of Biomarker Combinations in Fragile X Syndrome-Specific Clinical Trials: Evaluating Treatment Efficacy via Exploratory Biomarkers

**DOI:** 10.1101/2025.05.29.25328581

**Authors:** Jordan E. Norris, Elizabeth M. Berry-Kravis, Mark D. Harnett, Scott A. Reines, Melody A. Reese, Emma K. Auger, Abigail H. Outterson, Jeremiah Furman, Mark E. Gurney, Lauren E. Ethridge

## Abstract

Fragile X Syndrome (FXS) is a rare neurodevelopmental disorder caused by a trinucleotide repeat expansion on the 5’ untranslated region of the *FMR1* gene. FXS is characterized by intellectual disability, anxiety, sensory hypersensitivity, and difficulties with executive function. A recent phase 2 placebo-controlled clinical trial assessing BPN14770, a first-in-class phosphodiesterase 4D allosteric inhibitor, in 30 adult males (age 18-41 years) with FXS demonstrated cognitive improvements on the NIH Toolbox Cognitive Battery in domains related to language and caregiver reports of improvement in both daily functioning and language. However, individual physiological measures from electroencephalography (EEG) demonstrated only marginal significance for trial efficacy. A secondary analysis of resting state EEG data collected as part of the phase 2 clinical trial evaluating BPN14770 was conducted using a machine learning classification algorithm to classify trial conditions (i.e., baseline, drug, placebo) via linear EEG variable combinations. The algorithm identified a composite of peak alpha frequencies (PAF) across multiple brain regions as a potential biomarker demonstrating BPN14770 efficacy. Increased PAF from baseline was associated with drug but not placebo. Given the relationship between PAF and cognitive function among typically developed adults and those with intellectual disability, as well as previously reported reductions in alpha frequency and power in FXS, PAF represents a potential physiological measure of BPN14770 efficacy.

---

Fragile X Syndrome (FXS) is a rare X-linked, monogenic neurodevelopmental disorder caused by a trinucleotide repeat expansion of ≥ 200 CGG repeats in the 5’ untranslated region of the Fragile-X messenger ribonucleoprotein 1 (*FMR1*) gene resulting in gene methylation and a subsequent full or partial reduction in Fragile X messenger ribonucleoprotein (FMRP) output [1, 2]. The relationship between molecular disruptions and externally measurable cognitive and behavioral symptoms for FXS, and whether these are mediated by abnormal physiology, is unknown [3]. Identifying the relationship between molecular mechanisms and clinical measures indexing cognition is important for developing targeted treatments that address cognitive disruptions, including impaired verbal and nonverbal intelligence, language processing and production difficulties, crystalized cognition issues, and cognitive inflexibility [4, 5]. Cognitive symptoms and difficulties with learning are among the most distressing symptoms for individuals with FXS and their families as intellectual disability (ID) limits independence in activities of daily living [6, 7, 8, 9]. Current behavioral and pharmaceutical interventions aim to address symptoms of FXS but without specifically targeting the underlying pathophysiology of FXS, or the ID/cognitive deficits [10, 11].

Recent findings from a successful phase 2 clinical trial assessing the novel therapeutic BPN14770 (Zatolmilast) demonstrated good safety and tolerability with improvements on measures of cognition and daily functioning, showing promise as the first pharmaceutical intervention targeting the underlying pathophysiology of cognition in FXS [11]. BPN14770 is a first-in-class phosphodiesterase-4D (PDE4D) allosteric inhibitor, which is specific for the dimeric, PKA-activated form of PDE4D that acts as a key modulator of cAMP levels relevant to important cognitive functions, such as learning and memory (**Figure 1)**. Reduction of FMRP in FXS affects typical cAMP metabolism such that the decreases in cAMP are often observed in FXS [12, 13, 14]. While the phase 2 trial showed a cognitive benefit for BPN14770, understanding of the impact of BPN14770 on EEG-derived physiological measures remains incomplete.

**Figure 1.**
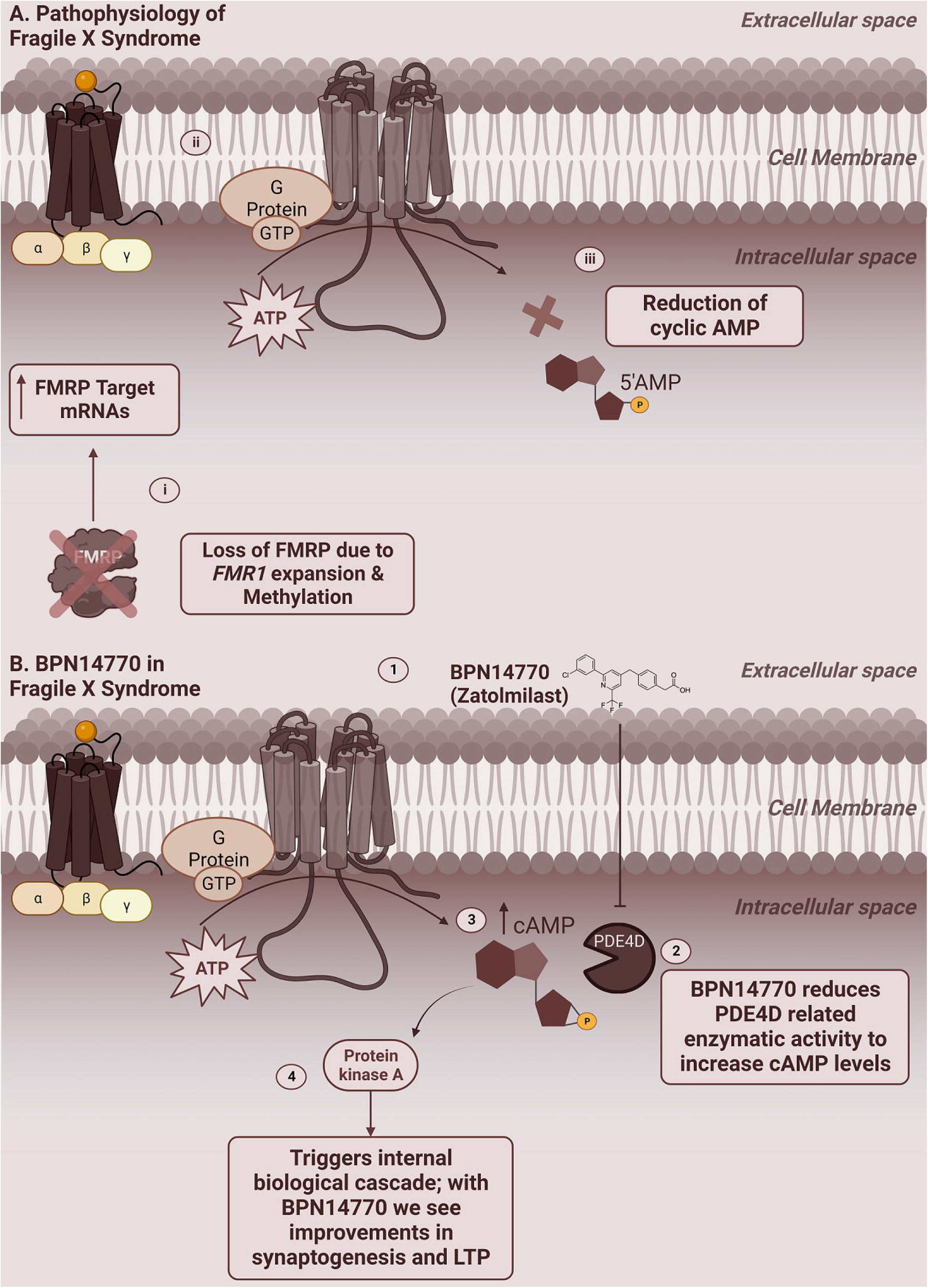
**A)** The pathophysiology of FXS: **i-iv.)** Reduced FMRP results in aberrant patterns of protein translation, internalization of AMPA receptors and reduced levels of cAMP. within neurons **B)** The mechanistic action of BNP14770: **1-4** BPN14770 inhibits activity of phosphodiesterase 4D or PDE4D leading to increased cAMP circulation within neurons resulting in improvements in synaptogenesis and LTP. Made with BioRender.

To determine the impact of BPN14770 on potential EEG biomarkers associated with clinical change, EEG was obtained in the phase 2 clinical trial. Specifically, N1 event-related potential (ERP) amplitude and habituation to repeated auditory stimulation were selected *a priori* for assessment based on previous findings of increased N1 amplitudes and decreased N1 habituation in FXS [15]. Differences between treatment and placebo were marginally significant (p = .06), and N1 amplitude was correlated with serum levels of drug suggesting some reduction in abnormally elevated N1 amplitude with BPN14770 [11, 16]. However, comparisons were underpowered due to data loss in the ERP, which is a longer and thus relatively more difficult-to-collect measure in FXS. Exploratory resting state EEG (rsEEG) was also collected to evaluate improvements in frequency bands most affected in FXS. rsEEG is a rich source of information about intrinsic neural activity and analyses of rsEEG data are generally more robust against data loss compared to the ERP. In previous rsEEG studies, individuals with FXS typically exhibit increased power in gamma (30 – 90 Hz) and theta (4 – 7 Hz) with notable decreases in alpha (8 – 13 Hz) bands [15, 17, 18], as well as reduced dynamic utilization of alpha oscillations, and a downshifted peak alpha frequency (PAF) which is known to correlate with cognitive performance in typically developed individuals [17, 18, 19, 20, 21, 22, 23]. Re-examination of EEG with a focus on rsEEG data may reveal underlying shifts associated with BPN14770 in biomarkers that index FXS pathophysiology.

Taking a more data-driven approach to evaluating the rsEEG may help reduce sample size concerns when exploring physiological correlates of BPN14770 efficacy. Neural biomarker development via EEG and translation of biomarkers to clinical-based interventions in FXS research supports efforts to identify novel therapeutic interventions specific to cognitive outcomes in FXS. However, data loss in populations with FXS presents a challenge to determining therapeutic efficacy via decreases in statistical power, particularly in phase 1 and 2 studies which typically recruit minimally necessary samples to establish safety and tolerability. Despite a focus on establishing safety and tolerability in phase 2 studies, determination of efficacy and target engagement is still required to move into phase 3. Efficacy challenges faced by clinical trials can often reflect methodological difficulties rather than a definitive lack of target engagement. Approximately 60% of clinical trials involving novel therapeutics that fail in clinical development, fail due to inadequate evidence for efficacy where inability to demonstrate efficacy may occur due to misspecification of the best endpoint, making secondary analysis an important follow-up step [20]. Given the demonstration of efficacy in cognitive measures in the current phase 2 clinical trial assessing BPN14770, concomitant physiological changes were likely present but not as robust in EEG due to a mismatch in the chosen endpoint (N1 amplitude), statistical underpowering due to data loss in the ERP task, or by focus on singular rather than composite EEG measures [11]. Composite measures, which integrate patterns across multiple variables, may provide optimal protection from chance fluctuation in single variables, thus increasing signal-to-noise allowing better capture of physiological change in small samples [21]. While biomarkers are excellent tools for improving diagnostic specificity, measuring therapeutic efficacy, and mechanistically understanding biological processes, utilizing single biomarkers may not fully characterize physiological dynamics relevant to detection of effects in the smaller samples typical of phase 2 clinical trials. Recent work in FXS and other populations has demonstrated that combining biomarkers to index nuanced physiological patterns may allow for more robust measurement at the individual level that will enhance relationships to clinically relevant outcomes (i.e., changes in observable behaviors) and be robust against consequences of data loss for individual measures [25, 26, 27]. However, introducing multiple combinations of measures for statistical comparison can lead to multiple comparisons concerns and increase chance of Type I error, necessitating some data reduction strategy to select the most likely variable combinations in advance this step. Machine learning classifiers, most frequently used to identify and separate diagnostic groups via receiver operating curve (ROC) analyses [28], may also be utilized to identify and separate treatment conditions, and can serve as an important screening step for composite biomarker identification.

The current study constitutes a secondary analysis of rsEEG using data driven methods to explore overarching shifts in neural physiology by utilizing a naïve Bayes Classifier to determine whether *a priori* linear variable combinations can separate participants along trial conditions (i.e., pre-dose baseline, placebo, and BPN14770) for the phase 2 trial of BPN14770 in FXS. By evaluating the feasibility of utilizing machine learning to ”pre-screen” composite variable biomarkers in this clinical trial with limited sample size, we aim to further address target engagement and physiological effects of BPN14770 on neural processes of interest in FXS.

## Methods

### Study Design

Participants were 30 males (full sample: age 18-41 years, *M* = 31.63, *SD* = 7.32; IQ 24.63-66.19, *M* = 42.78, *SD* = 11.16) with FXS participating in a single-center, phase 2a clinical trial assessing the efficacy and safety of BPN14770. The clinical trial was randomized, double-blinded, placebo-controlled, and utilized a two-period cross-over design without a washout period (**Figure 2**; ClinicalTrials.gov identifier: NCT03569631, see [11] for full inclusion and exclusion criteria). All participants or their legal guardians signed informed consent which included consent for EEG data collection. The clinical site was Rush University Medical Center (RUMC) where all study documents, including study protocol, consent documents, recruitment materials, safety information for participants, and information about study compensation were approved by the RUMC institutional review board [11]. Participant recruitment was managed by RUMC and supplemented by FXS patient advocacy groups. All EEG procedures were approved by the University of Oklahoma institutional review board.

**Figure 2.**
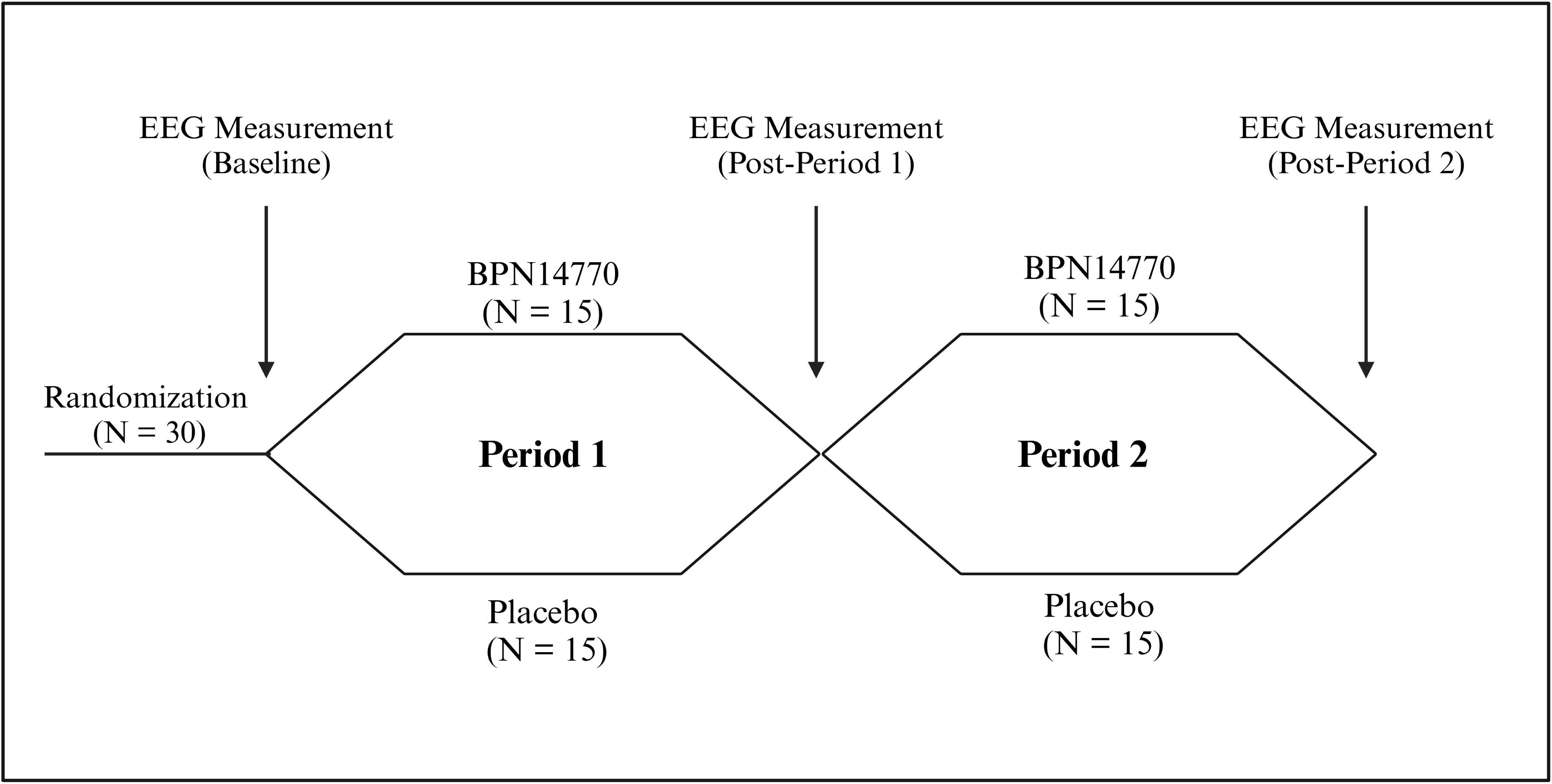
Flow Diagram for Clinical Trial Design with Indication of EEG Measurement Timepoints. The phase 2a trial evaluating BPN14770 (Zatomilast) utilized a cross-over design and enrolled 30 individuals with FXS. The 30 were then randomized into two groups and the first (baseline) EEG measurement was taken during screening. During period 1, half received BPN14770 and the other group received placebo. After period 1, another EEG was taken and was either the drug condition or placebo condition EEG measure depending on group. During period 2, the groups switched, and they received whichever intervention conditions they had not received in period 1. After period 2, the final EEG measurement was taken. Made with BioRender.

The current methods cover a secondary analysis of rsEEG data collected to explore biomarker outcomes for assessing target engagement. There was some initial EEG data loss in period 2 due to the study being completed partly during the Covid19 pandemic. The baseline sample consisted of 23 individuals. Of the 23, 17 provided quality EEG data across all conditions for both periods (i.e., BPN14470 and placebo). Two additional individuals were missing a drug or placebo measurement where multiple imputation was used to impute the relationship between drug and baseline for statistical comparison only (all conditions sample: N = 19 ages 21-41 years; *M* = 32.95, *SD* = 6.58; IQ 24.63-63.41, *M* = 41.20, *SD* = 10.37). Finally, the period 1 sample consisted of 23 who provided quality placebo or BPN14770 EEG data for period 1 and were used to evaluate BPN14770 against placebo as a between subjects’ comparison (N = 23, age 21-41 years, *M* = 32.74, *SD* = 6.57; IQ 24.63-63.41, *M* = 41.46, *SD* = 9.44).

### EEG Recording and Preprocessing

EEG data were continuously recorded and digitized at 512Hz, with a 5th order Bessel anti-aliasing filter at 200 Hz, using a 32-channel BioSemi ActiveTwo system (BioSemi). All sensors were referenced to a Common Mode Sense-Driven Right Leg active reference loop which replaces traditional ground electrodes and actively corrects for electrical noise during recording. Data were inspected offline and preprocessed to remove artifacts prior to analysis. No more than ∼ 5% of sensors were interpolated (interpolation limited to a max of 2 channels); data were digitally filtered offline from 0.5 to 100 Hz with a 57-63 Hz notch, resampled to a 500 Hz sampling rate, submitted to independent components analysis (ICA) via EEGLAB for artifact removal with segments of data containing large movement-related artifacts that would negatively impact the ICA decomposition removed prior to ICA [29], and re-referenced to the average of all channels (see supplemental figure 1). Final data lengths were all greater than 20s for analysis (average data length in seconds: baseline: M = 92.04, SD = 39.56; BPN14770: M = 85.83, SD = 41.79; Placebo: M = 91.51, SD = 35.24). A repeated measures ANOVA was run to assess differences in data length across conditions (N = 19) with no main effect of data length, *F*(2,17) = .83, *p* = .45, ES = .09.

### Frequency Bands

Continuous absolute and relative power were calculated across all electrodes for each frequency band where spectral power density was divided into 7 bands: delta (2 – 3.5 Hz), theta (3.5 – 7.5 Hz), alpha 1 (8 – 10 Hz), alpha 2 (10 – 12.5 Hz), beta (13 – 30 Hz), gamma 1 (30 – 55 Hz), and gamma 2 (65 – 90 Hz) [17]. Power was calculated using the first 80 seconds of data to standardize across participants; if 80 seconds of data was not available the maximum amount of data available was used.

### Peak Alpha

Peak alpha frequency (PAF) was included as a biomarker of cognitive function. Frequency bins (0.5 Hz) were formed to create a power density spectrum (PDS) calculated from absolute power. The absolute power spectrogram was converted to a relative power spectrogram and log transformed to find the maximum peak between 6 and 14 Hz for each electrode from a given participant using methods consistent with previous work in FXS [17] (**Figure 3**; see supplemental figure 2).

**Figure 3.**
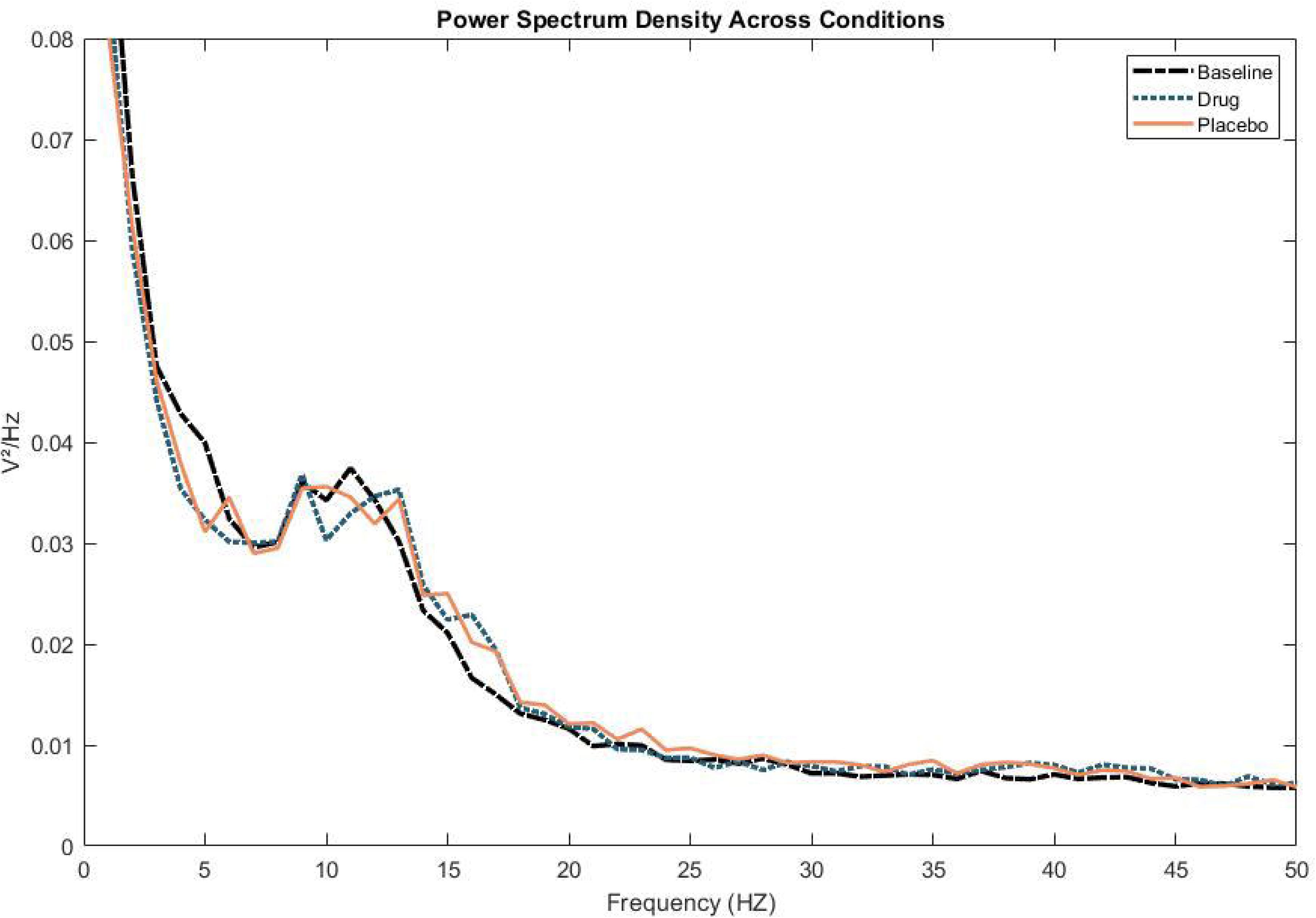
Relative Power Spectrum Density Plot Across Conditions. PSD for all conditions generated and used to compute PAF values.

### Alpha bursts

Neurodynamic metrics in both the alpha and gamma frequency bands were calculated. Methods for calculating alpha burst metrics reflect those proposed by Allen and Cohen (2010) and used by our group previously [20, 30]. Data were bandpass filtered in the alpha range (8 – 13 Hz) and a Hilbert transform was applied. The transformed signal was used to calculate instantaneous power at each timepoint using the natural log of the squared absolute value of the complex result. Burst metrics were calculated by selecting one *a priori* electrode from each hemisphere and subtracting left hemisphere data (i.e., F3/O1) from the right hemisphere data (i.e., F4/O2) to provide continuous asymmetry power values - [power(Right) – power(Left)]. Alpha bursts were then calculated by defining a threshold as the upper 80^th^ percentile of the continuous asymmetry power value for each participant where burst counts totaled the number of times the threshold value was crossed per second and lengths reflected the average time spent in each burst (i.e., continuous time spent above threshold). Alpha burst metrics were calculated from both frontal (i.e., F3/F4) and occipital (i.e., O1/O2) regions and gamma bursts were calculated only from the frontal region to avoid areas generating increased muscle artifact. Gamma burst metrics were calculated similarly to alpha burst metrics with the threshold set at the 90^th^ percentile to take a more conservative approach [20, 30].

### Biomarker Combinations

Predictors were hypothesis driven combinations of EEG measures previously reported to be affected in FXS. All biomarkers were EEG frequency measures evaluated across multiple metrics using different a priori electrode combinations. Metrics were assessed in the frontal region (i.e., electrodes FP1, FP2, AF3, AF4, F3, F4, F7, F7), occipital region (i.e., electrodes PO3, PO4, O1, O2), and across the whole head using all 32 electrodes. Broadly, predictors were pulled from three categories: 1.) frequency bands: individual frequency bands from combined frontal/occipital regions, specific frequency band combinations (e.g., alpha and theta), and region-specific assessments of all frequency bands across brain regions (frontal, occipital, and whole head), 2.) peak alpha across the same brain regions, and 3.) burst metrics.

### Biomarker Combinations: Frequency Bands

Relative and absolute power were computed across each frequency band and assessed from the combination of frontal and occipital regions due to targeted interest in frequency utilization in frontal and occipital regions. Combinations of specific frequency bands of interest were also assessed, including frontal theta-alpha, frontal theta-gamma, occipital theta-alpha, and theta-alpha-gamma combinations.

Lastly, all frequency bands calculated from both relative and absolute power (i.e., delta (2-3.5 Hz), theta (3.5-7.5 Hz), alpha 1 (8-10 Hz), alpha 2 (10-12.5 Hz), beta (13-30 Hz), gamma 1 (30-55 Hz), and gamma 2 (65-90 Hz)) were evaluated across frontal and occipital regions to assess region-specific frequency band effects, and then evaluated across the whole head.

### Biomarker Combinations: Peak Alpha

PAF predictor variable combinations were assessed by region (i.e., frontal, occipital, whole head, all three regions combined, and frontal/occipital combined). The frontal/occipital combination served as a more targeted assessment of region-specific differences in alpha frequency utilization, as differences in PAF have been reported between these regions in FXS [17], whereas whole head assessments provide a more parsimonious but less dynamic PAF measure.

### Biomarker Combinations: Burst Metrics

Burst metrics were included as an exploratory predictor set to explore whether BPN14770 improved dynamic alpha and gamma utilization. Alpha and gamma bursts were assessed in frontal (F3 and F4) and occipital (O1 and O2) regions with predictors reflecting length of time spent or number of times per second (CPS) individuals entered a dynamic alpha/gamma state. Specifically, predictors were: 1.) combination of all variables (F3/F4 CPS and lengths, O1/O2 CPS and lengths), 2.) lengths and CPS at specific electrodes (F3, F4, O1, and O2), or 3.) individual CPS or lengths within respective electrode sets (F3/F4, O1/O2). Gamma bursts were assessed for the frontal region only (F3 and F4) with predictors generated similarly to alpha burst metric predictors due to high muscle artifact from the neck region.

### Machine Learning Algorithm

A multiclass naïve Bayes classifier (NBC, see Supplemental Figure 3 for methodological details), selected for robustness against small sample sizes [31, 32] was used to classify participants into BPN14770, placebo, or baseline conditions based on hypothesis-driven variable combinations using MATLAB R2020b (The Mathworks, Natick, MA, United States). Code is available upon request to corresponding author. The NBC produces simple linear outputs that are easily translatable to clinical threshold values and calculation of composite scores. Importantly, the NBC performed on all three study conditions (baseline, placebo, drug) assumes that placebo is separable from baseline and thus that a placebo effect of some measurable magnitude occurs. We first evaluated all three study conditions on the 17 individuals that had complete usable data to evaluate physiological features robust against placebo effects and potential carryover effects (persistent pharmaceutical effects into the placebo window). Additional classifications were made following the same procedure due to persistent BPN14770 carryover effects in period 2 for those in the placebo condition: the NBC was additionally used to 1.) classify participants into placebo or BPN14770 for period 1 only, and 2.) baseline or BPN14770 across both period 1 and 2. The naïve Bayes model was trained on predictors using 70% of the data and tested on a holdout sample of 30%. Cross validated classification error (CE) was calculated for each combination using kfoldLoss to assess errors in correctly classifying the whole dataset. Due to the limited sample size, the process was bootstrapped over 2000 iterations to produce an overall average CE, area under the curve (AUC) for the ROC, true positive rate, and false positive rate for assessing model performance. Statistical evaluations were limited to a subset of variable combinations that performed best per AUC and CE values.

### Clinical Measures

Clinical trial secondary outcome measures assessed in the current study included the 1) National Institutes of Health-Toolbox Cognition Battery (NIH-TCB) which included 5 subscales (i.e., Cognition Crystallized Composite (CCC), Picture Vocabulary (PV), Oral Reading Recognition (ORR), Picture Sequence Memory (PSM), and Pattern Comparison Processing Speed PCPS) [33], 2) a Visual Analog Scale constructed using patient-specific behavioral anchors selected by the parent/caregiver to assess 3 domains (language, anxiety/irritability, and daily function) [11], 3) Aberrant Behavioral Checklist [34], and 4) Anxiety, Depression, and Mood Scale [35]. The original study found significant improvements with BPN14770 in NIH-TCB CCC, PV, ORR, and both VAS daily functioning and language domains. Clinical variables were selected because they either 1) demonstrated BPN14770 effects or, 2) are related to processes mediated by PAF.

### Statistics

Difference scores (i.e., either baseline or placebo effects subtracted from BPN14770 effects) were calculated to match NBC methods for generating probabilities for statistical evaluation of BPN14770 effects. Difference scores were evaluated using one-sided one-sample t-tests based on expected performance in favor of BPN14770. One-sided tests were assessed because statistics were only applied to best-performing variables. Repeated measure ANOVAs assessed differences on best performing EEG variable combinations and clinical variables across conditions with Fisher’s LSD to assess for condition differences when significant main effects were present. Effect sizes are reported as partial eta squared. Linear regressions were used to assess causal relationships between best performing variables/variable combinations and clinical variables that previously detected BPN14770 effects [11]. We examined exploratory correlations between best performing EEG variables and clinical variables for both BNP14770, placebo, and baseline using Spearman’s rho. Baseline correlations were evaluated to establish relationships between processes captured by clinical measures and EEG to support validity of any clinical correlations with EEG for the BPN14770 condition. Further, baseline correlations were an exploratory effort to better establish relationships between EEG measures and clinical features of FXS. For period 1 only, independent samples t-tests were conducted to assess differences between placebo and BPN14770. Effect sizes for all t-test are reported as Cohen’s d.

## Results

### Naïve Bayes Classifier Performance

150 hypothesis-driven variable combinations per comparison (i.e., all conditions, period 1, and baseline vs. drug) were evaluated by the NBC, for a total of 450 total combinations. PAF was identified as the best performing variable category with the combination of all variables outperforming single regions or other region combinations determined by which variable had the highest AUCs and lowest CE (**Table 1**). PAF was evaluated statistically for neural effects in favor of BPN14770 across all conditions (N = 19) and in period 1 only (N = 23) due to known carryover effects.

**Table 1.**
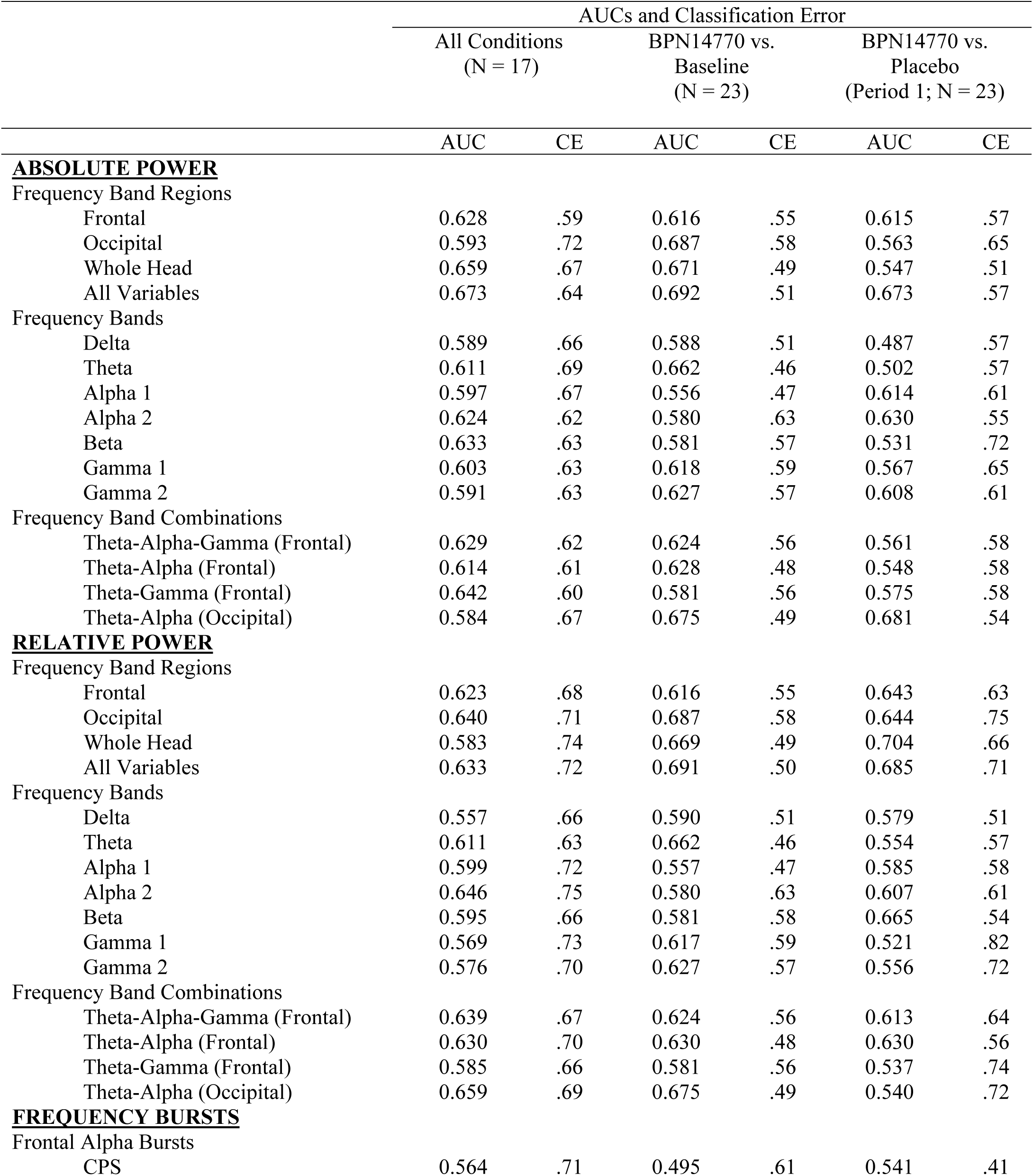
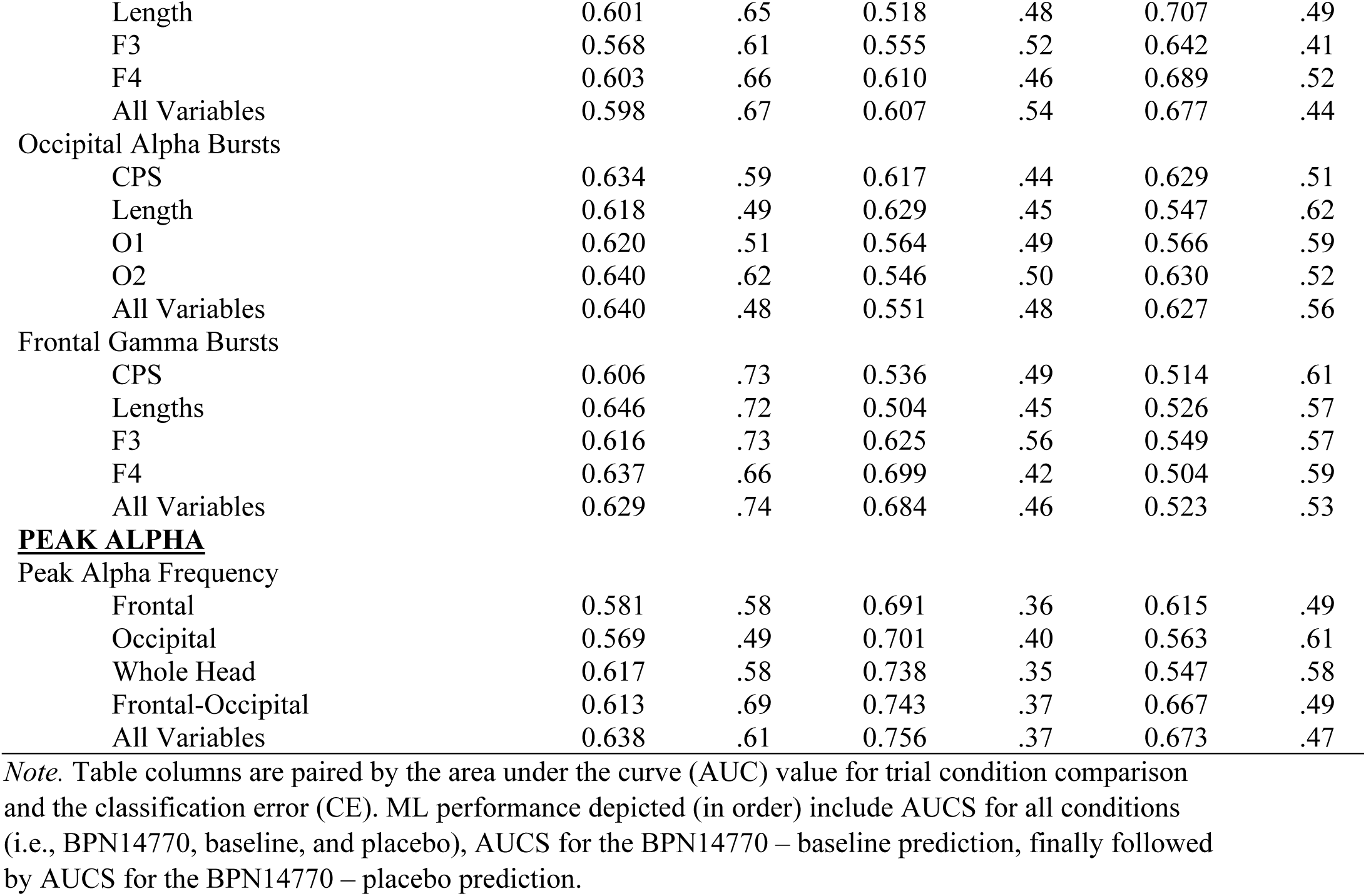
Area Under the Curve (AUC) and Classification Errors for all Variable Combinations.

|  |  | AUCs and Classification Error |  |  |  |  |  |
| --- | --- | --- | --- | --- | --- | --- | --- |
|  |  | All Conditions<br>(N = 17) |  | BPN14770 vs.<br>Baseline<br>(N = 23) |  | BPN14770 vs.<br>Placebo<br>(Period 1; N = 23) |  |
|  |  | AUC | CE | AUC | CE | AUC | CE |
| **<u>ABSOLUTE POWER</u>** |  |  |  |  |  |  |  |
| Frequency Band Regions |  |  |  |  |  |  |  |
|  | Frontal | 0.628 | .59 | 0.616 | .55 | 0.615 | .57 |
|  | Occipital | 0.593 | .72 | 0.687 | .58 | 0.563 | .65 |
|  | Whole Head | 0.659 | .67 | 0.671 | .49 | 0.547 | .51 |
|  | All Variables | 0.673 | .64 | 0.692 | .51 | 0.673 | .57 |
| Frequency Bands |  |  |  |  |  |  |  |
|  | Delta | 0.589 | .66 | 0.588 | .51 | 0.487 | .57 |
|  | Theta | 0.611 | .69 | 0.662 | .46 | 0.502 | .57 |
|  | Alpha 1 | 0.597 | .67 | 0.556 | .47 | 0.614 | .61 |
|  | Alpha 2 | 0.624 | .62 | 0.580 | .63 | 0.630 | .55 |
|  | Beta | 0.633 | .63 | 0.581 | .57 | 0.531 | .72 |
|  | Gamma 1 | 0.603 | .63 | 0.618 | .59 | 0.567 | .65 |
|  | Gamma 2 | 0.591 | .63 | 0.627 | .57 | 0.608 | .61 |
| Frequency Band Combinations |  |  |  |  |  |  |  |
|  | Theta-Alpha-Gamma (Frontal) | 0.629 | .62 | 0.624 | .56 | 0.561 | .58 |
|  | Theta-Alpha (Frontal) | 0.614 | .61 | 0.628 | .48 | 0.548 | .58 |
|  | Theta-Gamma (Frontal) | 0.642 | .60 | 0.581 | .56 | 0.575 | .58 |
|  | Theta-Alpha (Occipital) | 0.584 | .67 | 0.675 | .49 | 0.681 | .54 |
| **<u>RELATIVE POWER</u>** |  |  |  |  |  |  |  |
| Frequency Band Regions |  |  |  |  |  |  |  |
|  | Frontal | 0.623 | .68 | 0.616 | .55 | 0.643 | .63 |
|  | Occipital | 0.640 | .71 | 0.687 | .58 | 0.644 | .75 |
|  | Whole Head | 0.583 | .74 | 0.669 | .49 | 0.704 | .66 |
|  | All Variables | 0.633 | .72 | 0.691 | .50 | 0.685 | .71 |
| Frequency Bands |  |  |  |  |  |  |  |
|  | Delta | 0.557 | .66 | 0.590 | .51 | 0.579 | .51 |
|  | Theta | 0.611 | .63 | 0.662 | .46 | 0.554 | .57 |
|  | Alpha 1 | 0.599 | .72 | 0.557 | .47 | 0.585 | .58 |
|  | Alpha 2 | 0.646 | .75 | 0.580 | .63 | 0.607 | .61 |
|  | Beta | 0.595 | .66 | 0.581 | .58 | 0.665 | .54 |
|  | Gamma 1 | 0.569 | .73 | 0.617 | .59 | 0.521 | .82 |
|  | Gamma 2 | 0.576 | .70 | 0.627 | .57 | 0.556 | .72 |
| Frequency Band Combinations |  |  |  |  |  |  |  |
|  | Theta-Alpha-Gamma (Frontal) | 0.639 | .67 | 0.624 | .56 | 0.613 | .64 |
|  | Theta-Alpha (Frontal) | 0.630 | .70 | 0.630 | .48 | 0.630 | .56 |
|  | Theta-Gamma (Frontal) | 0.585 | .66 | 0.581 | .56 | 0.537 | .74 |
|  | Theta-Alpha (Occipital) | 0.659 | .69 | 0.675 | .49 | 0.540 | .72 |
| **<u>FREQUENCY BURSTS</u>** |  |  |  |  |  |  |  |
| Frontal Alpha Bursts |  |  |  |  |  |  |  |
|  | CPS | 0.564 | .71 | 0.495 | .61 | 0.541 | .41 |

|  |  |  |  |  |  |  |
| --- | --- | --- | --- | --- | --- | --- |
| Length | 0.601 | .65 | 0.518 | .48 | 0.707 | .49 |
| F3 | 0.568 | .61 | 0.555 | .52 | 0.642 | .41 |
| F4 | 0.603 | .66 | 0.610 | .46 | 0.689 | .52 |
| All Variables | 0.598 | .67 | 0.607 | .54 | 0.677 | .44 |
| Occipital Alpha Bursts |  |  |  |  |  |  |
| CPS | 0.634 | .59 | 0.617 | .44 | 0.629 | .51 |
| Length | 0.618 | .49 | 0.629 | .45 | 0.547 | .62 |
| O1 | 0.620 | .51 | 0.564 | .49 | 0.566 | .59 |
| O2 | 0.640 | .62 | 0.546 | .50 | 0.630 | .52 |
| All Variables | 0.640 | .48 | 0.551 | .48 | 0.627 | .56 |
| Frontal Gamma Bursts |  |  |  |  |  |  |
| CPS | 0.606 | .73 | 0.536 | .49 | 0.514 | .61 |
| Lengths | 0.646 | .72 | 0.504 | .45 | 0.526 | .57 |
| F3 | 0.616 | .73 | 0.625 | .56 | 0.549 | .57 |
| F4 | 0.637 | .66 | 0.699 | .42 | 0.504 | .59 |
| All Variables | 0.629 | .74 | 0.684 | .46 | 0.523 | .53 |
| **<u>PEAK ALPHA</u>** |  |  |  |  |  |  |
| Peak Alpha Frequency |  |  |  |  |  |  |
| Frontal | 0.581 | .58 | 0.691 | .36 | 0.615 | .49 |
| Occipital | 0.569 | .49 | 0.701 | .40 | 0.563 | .61 |
| Whole Head | 0.617 | .58 | 0.738 | .35 | 0.547 | .58 |
| Frontal-Occipital | 0.613 | .69 | 0.743 | .37 | 0.667 | .49 |
| All Variables | 0.638 | .61 | 0.756 | .37 | 0.673 | .47 |

*Note.* Table columns are paired by the area under the curve (AUC) value for trial condition comparison and the classification error (CE). ML performance depicted (in order) include AUCS for all conditions (i.e., BPN14770, baseline, and placebo), AUCS for the BPN14770 – baseline prediction, finally followed by AUCS for the BPN14770 – placebo prediction.

## Statistical Evaluation of NBC Best Performers

### All Conditions (N = 19)

The best performing variable/variable combination was all PAF variables combined (**Figure 4A & 4B**). The average difference score for all variables combined assessing the BPN14470 effect minus pre-dose baseline was significantly different from 0, *t*(18) = 3.53, *p* = .001, d = .81. Across all conditions individually, BPN14770 effect minus pre-dose baseline PAF from frontal, occipital, and whole head regions were all significantly different from 0 (frontal: *t*(18) = 2.34, *p* = .031, d = .54; occipital: *t*(18) = 2.62, *p* = .017, d = .60; whole head: *t*(18) = 4.01, *p* = .001, d = .92). The average difference score (i.e., all head regions) for the BPN14770 effect minus placebo was not significantly different from 0, *t*(18) = 1.29, *p* = .211, d = .30. BPN14770 effect minus placebo was not different from 0 across all regions for PAF (frontal: *t*(18) = 1.57, *p* = .134, d = .36; occipital: *t*(18) = .38, *p* = .708, d = .09; whole head: *t*(18) = 1.52, *p* = .145, d = .34).

**Figure 4.**
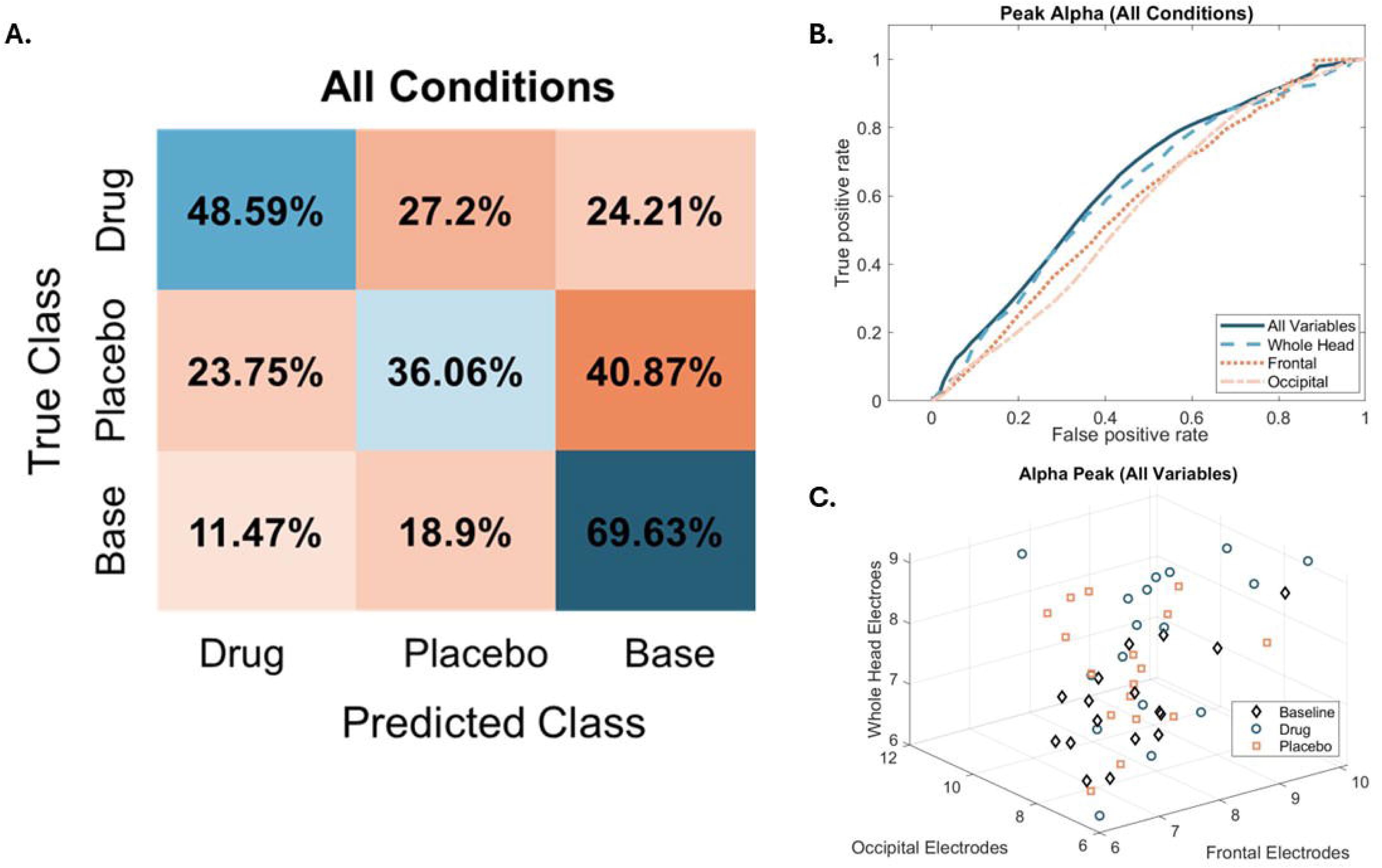
Average Classification Confusion Matrix Across All PAF Conditions, ROC curve, and Individual Data Points across All Conditions for all peak alpha. **A)** Average classification error confusion matrix showing classification performance in percentages of whether the true class was identified by the NBC. Percentage values reflect the average NBC performance across 2000 iterations of model building and across the 17 participants with data for all conditions. Misclassification is largely due to the overlap of placebo and baseline, but misclassification of drug and placebo can be attributed to carryover effects. **B)** ROC plot for PAF across head regions. Electrode selections included all electrodes (whole head), all frontal electrodes (frontal), all occipital electrodes (occipital), and all variables together (whole head, frontal, and occipital). Peaking toward the left upper corner indicates better performance (i.e., maximizing true positive rate and minimizing false positive rate). **C)** Individual data points for PAF across frontal, occipital, and all (i.e., whole head) electrodes.

A repeated measure ANOVA was conducted on PAF from all variables combined created by averaging PAF across all regions (i.e., frontal, occipital, and whole head) to assess for differences across conditions (**Figure 4**). There was a main effect of condition on PAF where BPN14770 increased PAF (*M* = 8.02, *SE* = 0.20) compared to baseline (*M* = 7.36, *SE* = 0.13) but not placebo (*M* = 7.71, *SE* = 0.17), *F*(2, 17) = 7.59, *p* = .004, ES = .47. Placebo represented an intermediate value, likely due to carryover effects from period 1, and was also significantly increased from baseline based on a post hoc comparison (p = .044).

The clinical variables were re-evaluated to determine whether significant clinical effects were detectable with reduced measurement time-points [i.e., 3 instead of the 5 reported in Berry-Kravis et al. (2021)] and reported in Supplemental Table 1 and 2. Correlations between PAF region difference scores (BPN14770-baseline) and difference scores for clinical variables of interest were assessed and found no significant relationships (Supplemental Table 3). Additionally, exploratory correlations were assessed for the BPN14770 condition only for PAF raw values (Supplemental Table 4) and found a significant correlation between whole head PAF and ABC lethargy/withdrawal, r = .53, p = .019. Another significant correlation was noted between frontal PAF and ADAMs obsessive/compulsive behavior, r = .46, p = .049. Other exploratory assessments between PAF and clinical variables were assessed and reported in supplement (Supplemental Table 5).

### Period 1 (N = 23)

BPN14770 differences from baseline were confirmed in period 1 with paired samples t-tests across all regions and found a similar pattern to that observed across both periods apart from occipital PAF. All between subject comparisons met criteria for equal variances across groups via Levene’s test. Frontal PAF in the BPN14770 condition (*M* = 7.69, *SD* = .90) was significantly different from baseline (*M* = 6.88, *SD* = .47), occipital PAF in the BPN14770 condition (*M* = 7.93, *SD* = 1.28) was not significantly different from baseline (*M* = 7.4, *SD* = .79), and whole head PAF in the BPN14770 condition (*M* = 7.79, *SD* = .75) trended towards a significance from baseline (*M* = 7.26, *SD* = .48), (frontal: *t*(11) = 2.53, *p* = .028, d = .73; occipital: *t*(11) = 1.18, *p* = .265, d = .34; whole head: *t*(11) = 2.08, *p* = .062, d = .60). Given the presence of carry-over effects, BPN14770 was tested against placebo in period 1. Independent sample t-tests were conducted on period 1 PAF data across frontal, occipital, and whole head regions and found no significant differences between BPN14770 and placebo (Frontal: t(21) = .302, *p* = .383, *d* = .126; Occipital: t(21) = .657, *p* = .259, *d* = .274; Whole head: t(21) = .833, *p* = .209, *d* = .348).

Paired samples t-tests were then conducted on period 1 PAF placebo and baseline data across all regions and found no significant differences between placebo and baseline except a marginal differences between baseline and placebo for occipital likely driven by a single individual (Frontal: t(8) = -.069, p = .947, d = - .023; Occipital: t(8) = 2.28, p = .052, d = .761; Whole head: t(8) = 1.75, p = .119, d =.526).

### Baseline Correlations (N = 24)

Exploratory correlations assessed relationships between pre-dose baseline PAF and pre-dose baseline clinical measures to determine the baseline relationships between PAF and clinical measures of interest (**Table 2**). A significant positive correlation was observed between frontal PAF and ABC Inappropriate Speech, *r* = .43, *p* = .036. A significant positive correlation was also observed between whole head PAF and ABC inappropriate speech (*r* = .46, *p* = .025), ABC stereotypy (*r* = .46, *p* = .025), and VAS daily functioning (*r* = .49, *p* = .015). Finally, a significant positive correlation was found between occipital PAF and ADAMs general anxiety (*r* = .41, *p* =.044), social anxiety (*r* = .42, *p*= .044), and manic and hyperactive behavior (*r* = .41, *p* = .040). Group differences for BPN14770 and placebo for clinical variables of interest are reported in Supplemental Table 6.

**Table 2.** Exploratory Correlations Assessing Pre-Dose Relationships between Peak Alpha Frequency and Clinical Measures.

| Variable (N) | Baseline Peak Alpha Frequency |  |  |
| --- | --- | --- | --- |
|  | Frontal PAF | Occipital PAF | Whole Head PAF |
| **NIH-TCB (N = 24)<sup>a</sup>** |  |  |  |
| CCC | -.120 | -.081 | .156 |
| PV | .064 | -.113 | .166 |
| ORR | -.286 | -.012 | .063 |
| PSM (N = 21) | -.103 | -.200 | .031 |
| PCPS | .159 | -.044 | -.155 |
| **VAS (N = 24)** |  |  |  |
| Language | .065 | -.177 | -.040 |
| Anxiety/Irritability | -.093 | .088 | .103 |
| Daily Functioning | .122 | .236 | .490* |
| **ABC (N = 24)** |  |  |  |
| Hyperactivity | .318 | .199 | .422* |
| Inappropriate Speech | .429* | .232 | .456* |
| Irritability | .176 | .094 | .266 |
| Lethargy/Withdrawal | .085 | .278 | .362 |
| Social Avoidance | -.017 | .092 | .206 |
| Stereotypy | .233 | .327 | .457* |
| **ADAMS (N = 24)** |  |  |  |
| Depressed Mood | -.127 | .219 | .089 |
| General Anxiety | .048 | .414* | .232 |
| Manic/Hyperactive Behavior | .170 | .422* | .278 |
| Obsessive/ Compulsive Behavior | .034 | .311 | .149 |
| Social Avoidance | -.086 | .415* | .266 |
| Deviation IQ (Z-score) | -.070 | -.417* |  |

*Note.* All correlations are Spearman's rho. Sample sizes are listed for each task and are the same across all sub-variables unless otherwise noted as N<sup>a</sup>. No asterisk = N.S., \*p < 0.05, \*\*p < 0.01.

## Discussion

The current study reflects an exploratory evaluation of secondary outcome measures from a recent, successful phase 2a clinical trial demonstrating BPN14770 efficacy in a small sample of males with FXS. The NBC identified PAF as a variable that adequately separated BPN14770 from baseline and placebo with the strongest predictor of condition being the combination of all PAF variables (frontal, occipital, and whole head PAF) which then demonstrated statistical significance for BPN14770 vs baseline but not placebo vs baseline. Our novel identification of BPN14770 efficacy in PAF adds to previous findings of cognitive improvements with BPN214770, as peak alpha is notably reduced in individuals with FXS and related to cognitive performance in typical development [17, 19, 36, 37].

### Improvements in PAF

Phase 2 clinical trials are frequently underpowered where multiple statistical comparisons across proposed biomarkers render secondary outcomes susceptible to false outcomes (i.e., type I and type II error). Utilization of classification algorithms represents a simplistic approach to secondary biomarker assessments in clinical trials. Not only are outcomes clinically interpretable and mechanistically insightful, where physiological outcomes help bridge known therapeutic mechanisms (i.e., pharmaceutical mechanisms of action) and externally measurable behavior/physiology, but the statistical evaluation is more robust against false discovery/type 1 error [21, 33]. Further, the current methods are easily employed under blinded conditions suggesting clinical trials in NDDs could move toward more data-driven outcome measures to avoid both type 1 and type 2 errors with more definitive efficacy determination.

Using the NBC to explore BPN14770-related physiological shifts identified adequate condition separation (i.e., baseline, drug, placebo) for PAF. Difficulty separating drug from placebo arose from known carryover effects which negatively affected NBC performance due to a lack of washout period in the cross-over design and persistent pharmacological effects in the placebo [11, 16]. The combination of all PAF variables separated drug from baseline with an area under the curve in the fair performance range for clinical use which survived statistical comparison [39]. Importantly, NBC performance on the PAF variable combination was achieved with a very small sample size (N = 17) of participants with clean EEG data across all timepoints. Statistical evaluations showed significant improvements in PAF with BPN14770 where PAF shifted into the alpha frequency range for all participants but were limited to evaluations comparing differences in BPN14770 effect from baseline and not placebo due to underpowered comparisons and carryover effects. Evaluating BPN14770 effects from placebo was only possible in period 1 rather than within participant comparisons. Despite the statistical limitations, the effect size for the linear composite variable assessing BPN14770 vs. Baseline created from all conditions was large and the effect size for BPN14770 vs placebo during period 1 was medium sized suggesting a moderate effect of BPN14770 on PAF. Further, the moderate effect of BPN14770 was driven by frontal measures of PAF indicating the effect may be specific to the thalamocortical generator of alpha [17].

Shifts in PAF are relevant given the nature of the clinical improvements initially observed in the cognitive domain despite not sharing relationships with clinical variables that initially demonstrated improvements [11]. PAF is associated with cognitive performance in typically developed individuals and correlates highly with measures of cognitive performance in both idiopathic ASD and samples of FXS that include females [17, 40, 41]. Further, PAF is typically considered highly stable in typically developed younger and older adults with evidence showing PAF was not easily modified by cognitive interventions alone on a larger time scale [42]. While PAF can fluctuate on smaller timescales and with varying task demands, males with FXS demonstrate difficulties initiating alpha frequency oscillations on a dynamic scale suggesting any intraindividual shifts into the alpha frequency range across time may be meaningful [43]. The current neocortical hyperexcitability model of FXS includes increased power in gamma and theta with decreased power in alpha [18]. Increased N1 amplitude in the ERP to a novel/initial auditory stimulus are thought to reflect neural hyperexcitability in FXS and related to blood serum levels of BPN14770 [15, 16, 44]. Combining increases in PAF with marginal improvements in the N1 amplitude measured from frontocentral electrodes reported in the original article detailing the clinical trial findings, BPN14770 may enhance an individual with FXS’s ability to initiate alpha frequency and organize neural networks necessary for supporting the process of temporal integration of information underlying both sensory processing and cognitive performance in frontocentral brain regions [11, 20,43]. Ultimately, given the lack of improvement observed from placebo, the adequate but not excellent AUCs for separating conditions, and the sample size limitations on statistical power, more work and a replication are required to add support to the current conclusions and establish PAF changes as a biomarker of efficacy for BPN14770.

### Limitations and Conclusions

First, the original manuscript reported clinical findings before and after each treatment arm (i.e., 5 total measurements) but EEG was recorded only at the end of each treatment arm (i.e., 3 total measurements) [11]. Thus, the current study was limited to one measurement per treatment arm for both clinical and EEG evaluations (see Figure 2). Clinical measure outcome differences between the original and current study (see supplement) may reflect the use of only 3 measurement points in the current study where 5 were used in the original or the reduction in sample size from 30 with clinical outcomes to 17 with complete EEG outcomes. Second, carryover effects were a major limitation for evaluating differences between BPN14770 and placebo. Third, we were unable to include the same covariates from the original study (i.e., IQ and baseline measures) due to 1.) the use of baseline measures in the analyses, and 2.) the relationship between IQ and PAF. Fourth, the use of machine learning to separate conditions also assumes that baseline values for the sample do not overlap significantly with the drug sample values, and so is only appropriate for variables where a mean shift at the group level in values is expected. In circumstances where there is a large range of variability in baseline values, with some individuals showing minimal impairment that overlaps with improved scores with treatment for those with lower baseline values, the classifier technique may underperform. We selected input variables with known deficits at the group level for FXS, therefore despite some overlap in conditions (**Figure 4C**) we do not expect this limitation to be significantly problematic for our study. However, this is an important consideration for use of this technique in trials with larger ranges of baseline ability. Finally, the NBC has an assumption of independence of features and certain EEG measures are likely correlated (e.g., power across bands) in certain instances. Although there is debate on how impactful this assumption is, dependence of some features likely reduces the effectiveness of the ML approach [45]. However, one benefit of the NBC independence assumption is that it allows it to learn high dimensional features and achieve model fit with much smaller training sets than many other classifiers [46, 47.] Despite these limitations, this study demonstrates the possibility of screening large numbers of exploratory variable combinations using machine learning, ROC evaluation, and composite variable generation in small sample sizes to detect novel effects of treatment on brain physiology. The composite PAF variable identified is relevant to the significant cognitive outcomes for BPN14770, has been demonstrated to be impaired in FXS [17], and is more robust to data loss than ERP measures, thus more scalable to larger trials. Future work will be necessary to validate PAF in larger sample sizes to assess the extent to which BPN14770 modulates cognition-related biomarkers in FXS.

## Supporting information

Supplemental Methods and Results

## Data Availability

Data availability requests must be made to Shionogi & Company.

## Acknowledgements

We thank the patients and their families for participating in the clinical trial. Direct clinical costs were funded by the FRAXA Research Foundation. Access to and training on the NIH-TCB was obtained in association with work on HD076189 (David Hessl). Tetra Therapeutics provided drug product and funded trial administration and independent data analysis. NFXF Randi Hagerman Summer Scholar Award funded J.E.N and the secondary analysis for the current study.

