## Supplemental Methods and Results for "ROC Analysis of Biomarker Combinations in Fragile X Syndrome-Specific Clinical Trials: Evaluating Treatment Efficacy via Exploratory Biomarkers"

Supplemental Materials

### Methods

Supplemental Figure 1.

*EEG Data Processing Flow Diagram*

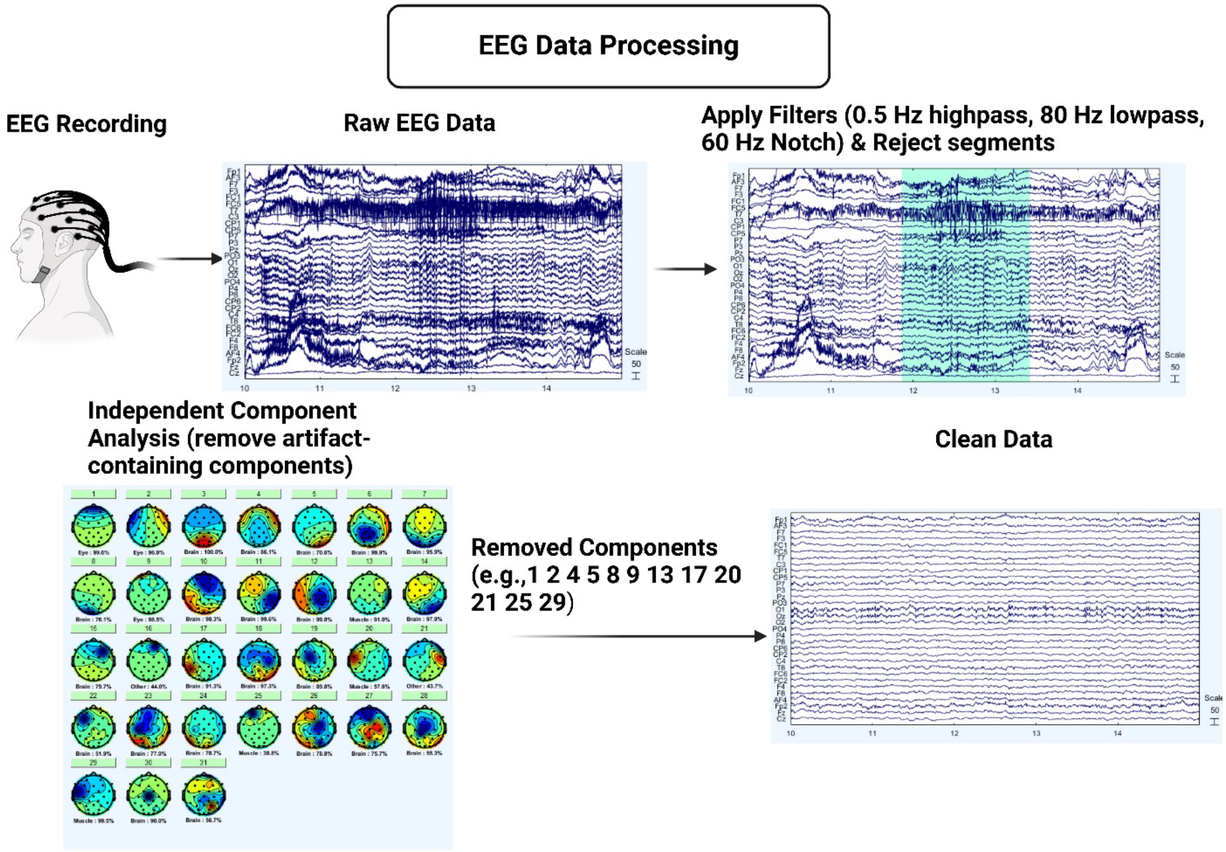


*Note.* The flow diagram depicts the process of first recording EEG data where both muscle and ocular artifacts are present in the electroencephalogram. Bad channels are interpolated (none present in examples). Artifacts are then removed either by rejection of swaths of data continuing artifact that will not decompose via independent component analysis (ICA). Data are then submitted to ICA for artifact-specific component removal. Made with BioRender.

**Supplemental Figure 2.**

*Data Analysis Flow Diagram*

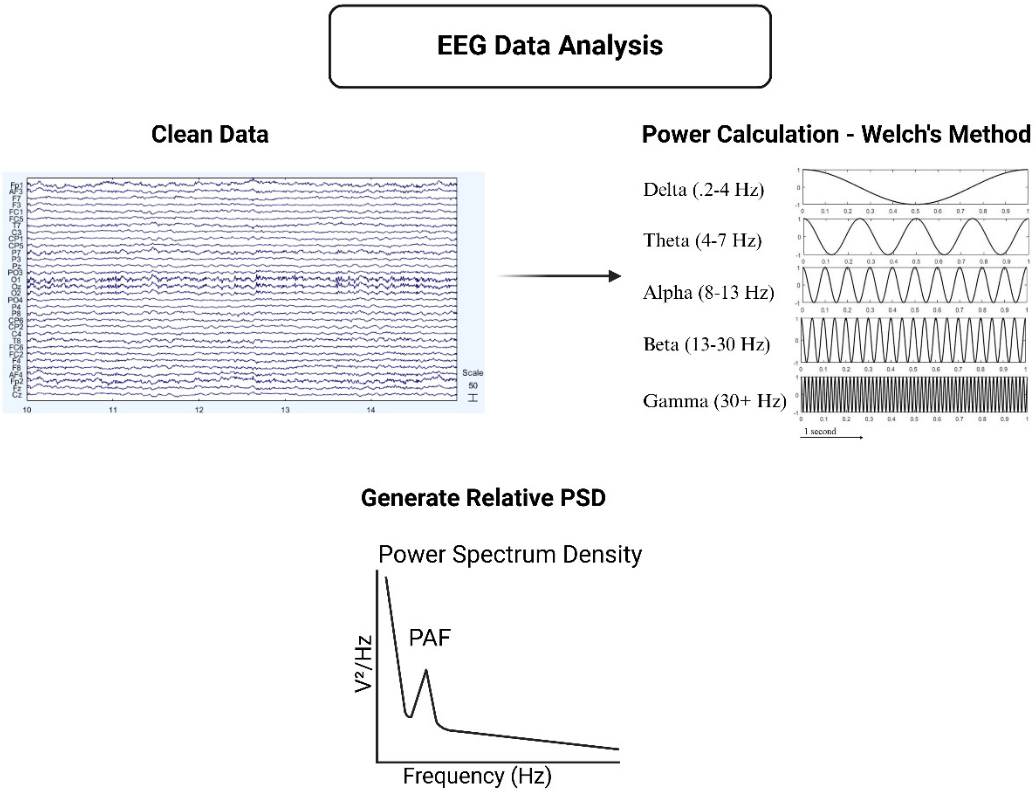


*Note.* Once the data are clean, power is computed using Welch's method which also generates the power spectrum density. Peak alpha frequency (PAF) is identified from the power spectrum density (PSD) using the MATLAB function 'findpeaks'. Of note, the power calculation and derivation of the PSD are relevant to all measurements except for the burst metrics which are described in the main methods and also in Norris et al. (2022). Made with BioRender.

Supplementary Figure 3.

Naïve Bayes Classifier (NBC) Flow Diagram

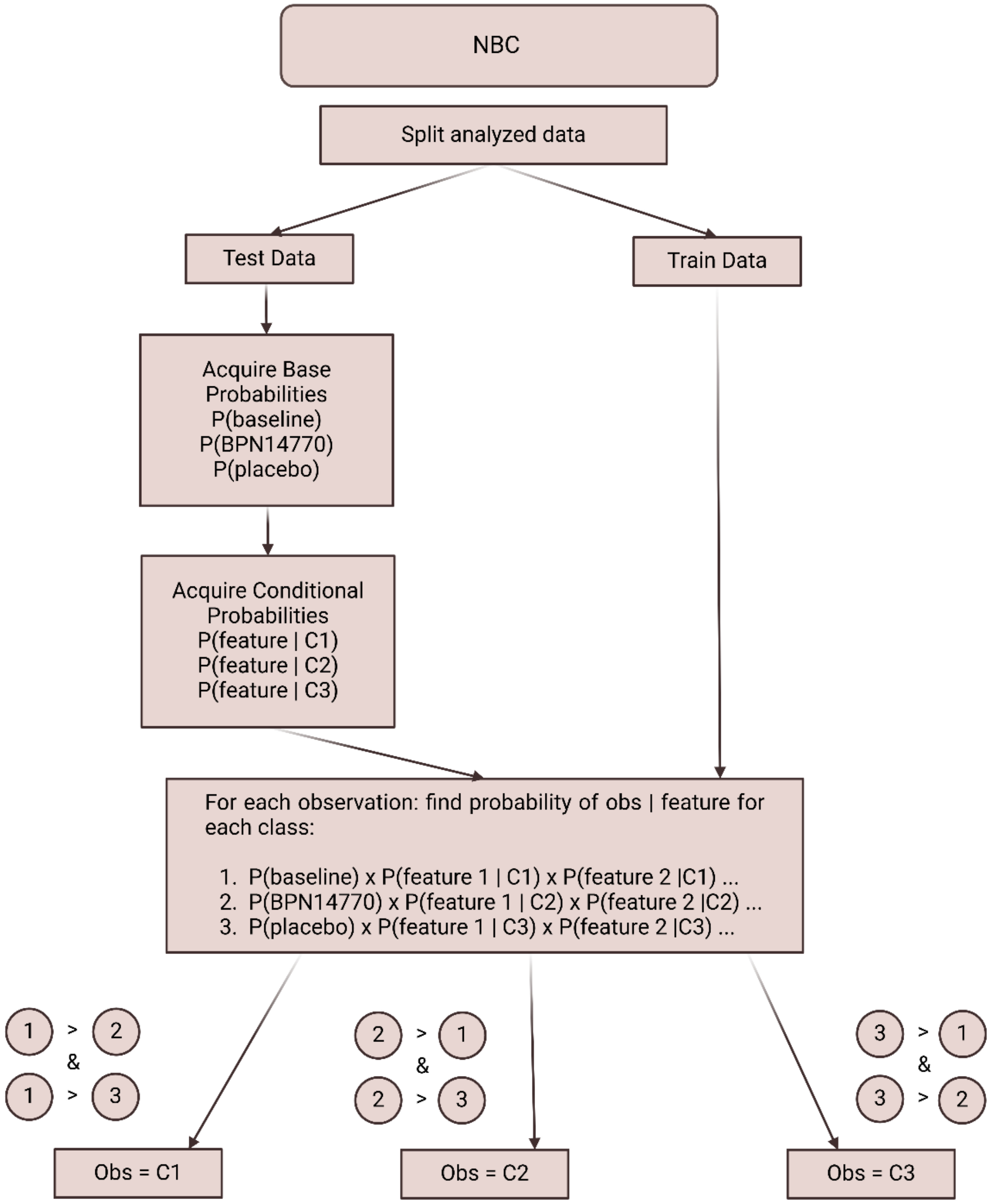

### Results

#### All Conditions (N = 19).

One-way rmANOVAs tested for differences across all conditions for clinical variables from the original study that showed improvements with BPN14770 (Supplemental Table 1). Only VAS language demonstrated significant improvements with BPN14770. There was a main effect of condition for VAS language demonstrating improvements with BPN14770 (M = 52.89, SE = 5.14) from baseline ($M$ = 33.11, $SE$ = 6.28) but not placebo ($M$ = 52.94, $SE$ = 6.53), $F(2, 17) = 8.27$, $p = .003$, ES = .49).

**Supplemental Table 1.**

*Repeated Measures ANOVA for Primary Clinical Variables*

| | rmANOVA Statistics | | | |
|---|---|---|---|---|
| Variable | df | f | p | ES |
| **NIH-TCB** | | | | |
| CCC | 2,17 | 0.24 | .788 | .03 |
| PV | 2,17 | 0.69 | .511 | .08 |
| ORR | 2,17 | 0.77 | .477 | .08 |
| PSM | 2,11 | 0.77 | .488 | .12 |
| PCPS | 2,17 | 1.66 | .221 | .16 |
| **VAS** | | | | |
| Language | 2,17 | 8.27 | .003 | .49 |
| Daily Functioning | 2,17 | 2.28 | .132 | .21 |

*Note.* The original study (Berry-Kravis et al., 2021) reported significant BPN14770 efficacy in the depicted variables. The variables were re-evaluated due to the reduced number of measurements included in the current study to determine whether clinical effects were still detectable with the use of measurements from only 3 time points to match EEG, as there were less EEG measurements (N = 3 time points) than clinical measurements (N = 5 time points). For visualization, see main manuscript Figure 2 and for more detail see the limitations section.

One-sample t-tests were conducted on clinical variable BPN14770-baseline difference scores for clinical variables that demonstrated BPN14770 effects in the original study to confirm BNP14770 effects (Supplemental Table 2). The only variable that demonstrated significant BPN14770 effects was the VAS Language domain, $t$ (18) = 4.129, $p$ < .001, d = .947. Both NIH-TCB PCPS and VAS anxiety/irritability trended towards significance (NIH-TCB PCPS: $t$(18) = 1.65, $p$ = .058, d = .379; VAS anxiety/irritability: $t$(18) = 1.518, $p$ = .073, d = .348).

**Supplemental Table 2.**

*One-sample t-tests assessing BPN14770 Effect in Clinical Variables of Interest.*

| | | | Statistics | | |
|---|---|---|---|---|---|
| **Clinical Variables** | t | df | p | d | 95% CI [LL, UP] |
| **NIH-TCB** | | | | | |
| CCC | -0.508 | 18 | .309 | -.117 | [-2.16, 1.32] |
| ORR | -1.240 | 18 | .116 | -.285 | [-1.84, .47] |
| PSM | 0.733 | 14 | .238 | .189 | [-5.52, 11.25] |
| PV | -0.063 | 18 | .475 | -.015 | [-3.59, 3.39] |
| **VAS** | | | | | |
| Language | 4.13 | 18 | .001 | .947 | [9.72, 29.86] |
| Anxiety/Irritability | 1.52 | 18 | .146 | .348 | [-3.52, 21.83] |
| Daily Functioning | 0.657 | 18 | .260 | .151 | [-9.61, 18.34] |

*Note.* Correlations assessed potential relationships between PAF variable BPN14770-baseline difference scores and clinical measure difference scores of interest but were not significant (Supplemental Table 3).

**Supplemental Table 3.**

*Correlations between PAF Difference Scores and Clinical Variables.*

| | BPN14770 – Baseline (N = 17) | | |
|---|---|---|---|
| **Variables (BPN14770 – baseline)** | | | |
| **NIH-TCB (N = 19)[a]** | **Frontal PAF** | **Occipital PAF** | **Whole Head PAF** |
| CCC | -.044 | -.103 | -.082 |
| PV | -.054 | -.126 | -.118 |
| ORR | -.134 | -.111 | -.158 |
| PSM **(N = 15)** | -.436 | -.323 | -.444 |
| PCPS | -.313 | -.031 | -.175 |
| **VAS (N = 19)** | | | |
| Language | .326 | -.041 | .162 |
| Anxiety/Irritability | .364 | -.164 | .129 |
| Daily Functioning | .309 | .197 | .241 |
| **ABC (N = 19)** | | | |
| Hyperactivity | .298 | -.098 | .304 |
| Inappropriate Speech | .216 | .689 | .206 |
| Irritability | .048 | .058 | .122 |
| Lethargy/Withdrawal | .323 | .104 | .296 |
| Social Avoidance | .181 | -.355 | .062 |
| Stereotypy | -.140 | -.213 | -.070 |
| **ADAMS (N = 19)** | | | |
| Depressed Mood | .125 | -.012 | .043 |
| General Anxiety | .059 | .255 | .117 |
| Manic/Hyperactive Behavior | -.016 | -.068 | -.080 |
| Obsessive/ Compulsive Behavior | -.318 | -.090 | -.379 |
| Social Avoidance | .140 | -.079 | .107 |

*Note*. All correlations are Spearman's rho. Sample sizes are listed for each task and are the same across all sub-variables unless otherwise noted as N[a]. No asterisk = N.S., *p <0.05, **p < 0.01.

Exploratory correlation analyses were conducted to explore relationships between raw PAF variables and all clinical variables for BPN14770 and placebo (Supplemental Table 4).

**Supplemental Table 4.**

*Exploratory Correlations Evaluating Relationships between Raw PAF Scores for BPN14770.*

| | | BPN14770 | |
|---|---|---|---|
| Variable | Frontal | Occipital | Whole head |
| **NIH-TCB (N = 19)[a]** | | | |
| CCC | -.418 | -.380 | -.445 |
| PV | -.300 | -.409 | -.395 |
| ORR | -.194 | -.107 | -.077 |
| PSM **(N = 16)** | -.413 | -.421 | -.387 |
| PCPS | -.158 | -.341 | -.084 |
| **VAS (N = 19)** | | | |
| Language | .020 | -.065 | -.016 |
| Anxiety/Irritability | .156 | -.054 | .074 |
| Daily Functioning | .081 | .087 | -.054 |
| **ABC (N = 19)** | | | |
| Hyperactivity | .051 | .035 | .075 |
| Inappropriate Speech | .183 | -.090 | .229 |
| Irritability | -.048 | .047 | .081 |
| Lethargy/Withdrawal | .422 | .230 | .534* |
| Social Avoidance | .194 | -.151 | .071 |
| Stereotypy | .373 | .097 | .332 |
| **ADAMs (N = 19)** | | | |
| Depressed Mood | .173 | -.160 | .145 |
| General Anxiety | .083 | .249 | .116 |
| Manic/Hyperactive Behavior | .052 | .065 | .149 |
| Obsessive/ Compulsive Behavior | .457* | .419 | .431 |
| Social Avoidance | .398 | .119 | .439 |

*Note*. All correlations are Spearman's rho. Sample sizes are listed for each task and are the same across all sub-variables unless otherwise noted as N[a]. No asterisk = N.S., *p <0.05, **p < 0.01.

Multiple linear regression models were assessed to evaluate causal relationships between PAF difference scores across all three head regions and clinical measure difference scores for clinical variables that demonstrated improvements with BPN14770 reported in the original study (Berry-Kravis et al., 2019) (Supplemental Table 5). No models were statistically significant but the model evaluating the BPN14770-placebo difference score across all head regions was marginal for NIH-TBC ORR with the BPN14770 effect explaining 38.6% of the variability in ORR scores ($F$(3,18) = 3.14, $p$ = .056, $R^2$ = .37), driven by the occipital region, β = -77, $t$(18) = -2.93, $p$ = .010.

**Supplemental Table 5.**

*Multiple Linear Regression Analysis Summary for Predicting BPN14770 Effect using Difference Scores.*

| Predictor | Coefficients | | | | ANOVA | | |
|---|---|---|---|---|---|---|---|
| | b | β | t | p | F | p | Fit ($R^2$) |
| **BPN14770 – Baseline** | | | | | | | |
| VAS Language | | | | | .66 | .587 | .12 |
| PAF Frontal | 9.38 | .60 | 1.33 | .202 | | | |
| PAF Occipital | 1.82 | .09 | 0.35 | .735 | | | |
| PAF Whole Head | -11.58 | -.41 | -0.87 | .398 | | | |
| VAS Daily Functioning | | | | | 2.27 | .122 | .31 |
| PAF Frontal | 21.35 | .98 | 2.48 | .026 | | | |
| PAF Occipital | 5.06 | .19 | 0.78 | .446 | | | |
| PAF Whole Head | -26.94 | -.69 | -1.65 | .119 | | | |
| NIH-TCB CCC | | | | | 0.21 | .207 | .04 |
| PAF Frontal | 0.76 | .28 | 0.59 | .559 | | | |
| PAF Occipital | 0.16 | .05 | 0.17 | .871 | | | |
| PAF Whole Head | -1.82 | -.37 | -0.76 | .461 | | | |
| NIH-TCB PV | | | | | 0.13 | .938 | .03 |
| PAF Frontal | 1.36 | .25 | 0.53 | .603 | | | |
| PAF Occipital | 0.58 | .09 | 0.30 | .768 | | | |
| PAF Whole Head | -3.07 | -.32 | -0.63 | .536 | | | |
| NIH-TCB ORR | | | | | 0.12 | .947 | .02 |
| PAF Frontal | 0.31 | .17 | 0.37 | .719 | | | |
| PAF Occipital | -0.15 | -.07 | -0.23 | .824 | | | |
| PAF Whole Head | -0.63 | -.19 | -0.39 | .701 | | | |
| NIH-TCB PSM | | | | | 1.56 | .255 | .29 |
| PAF Frontal | -3.67 | -.29 | -0.73 | .482 | | | |
| PAF Occipital | -4.58 | -.31 | -1.09 | .301 | | | |
| PAF Whole Head | -2.62 | -.11 | -0.25 | .808 | | | |
| NIH-TCB PCPS | | | | | 1.15 | .363 | .19 |
| PAF Frontal | -7.63 | -.77 | -1.79 | .094 | | | |
| PAF Occipital | -0.95 | -.08 | -0.29 | .771 | | | |
| PAF Whole Head | 10.22 | .58 | 1.27 | .224 | | | |
| **BPN14770 – Placebo** | | | | | | | |
| VAS Language | | | | | 1.72 | .205 | .26 |

| | | | | | | | |
|---|---|---|---|---|---|---|---|
| PAF Frontal | -8.05 | -.77 | -2.09 | .053 | | | |
| PAF Occipital | -3.11 | -.38 | -1.32 | .205 | | | |
| PAF Whole Head | 14.66 | .94 | 2.18 | .045 | | | |
| VAS Daily Functioning | | | | | 0.16 | .919 | .03 |
| PAF Frontal | -2.13 | -.18 | -0.42 | .680 | | | |
| PAF Occipital | -1.12 | -.12 | -0.36 | .722 | | | |
| PAF Whole Head | 5.96 | .33 | 0.67 | .512 | | | |
| NIH-TCB CCC | | | | | 0.64 | .602 | .11 |
| PAF Frontal | -1.03 | -.19 | -0.49 | .631 | | | |
| PAF Occipital | -1.73 | -.43 | -1.35 | .198 | | | |
| PAF Whole Head | 2.41 | .31 | 0.66 | .521 | | | |
| NIH-TCB PV | | | | | 0.38 | .767 | .07 |
| PAF Frontal | -1.67 | -.22 | -0.55 | .593 | | | |
| PAF Occipital | -1.11 | -.19 | -0.59 | .563 | | | |
| PAF Whole Head | 0.77 | .07 | 0.14 | .888 | | | |
| NIH-TCB ORR | | | | | 3.14 | .056 | .39 |
| PAF Frontal | -0.49 | -.12 | -0.36 | .724 | | | |
| PAF Occipital | -2.46 | -.77 | -2.93 | .010 | | | |
| PAF Whole Head | 3.73 | .61 | 1.55 | .142 | | | |
| NIH-TCB PSM | | | | | 2.07 | .167 | .38 |
| PAF Frontal | -3.78 | -.48 | -1.21 | .255 | | | |
| PAF Occipital | -1.30 | -.17 | -0.51 | .622 | | | |
| PAF Whole Head | -0.79 | -.07 | -0.14 | .891 | | | |
| NIH-TCB PCPS | | | | | 1.57 | .239 | .24 |
| PAF Frontal | -3.98 | -0.49 | -1.33 | .202 | | | |
| PAF Occipital | -3.74 | -0.60 | -2.05 | .059 | | | |
| PAF Whole Head | 7.39 | 0.62 | 1.41 | .178 | | | |

*Note*. All variables (i.e., PAF and clinical variables) are difference scores. Evaluation of placebo effects includes effects from period 2 which contained BPN14770 carryover effects.

**Period 1 (N = 23).**

Independent samples t-tests were conducted on clinical variables from period 1 only (Supplemental Table 6).

**Supplemental Table 6.**

*Independent Samples T-tests assessing BPN14770 vs. Placebo Effects for Clinical Variables of Interest*

| | BPN14770 | | Placebo | | Group Differences | | | |
|---|---|---|---|---|---|---|---|---|
| Variable | M | SD | M | SD | df | t | p | d |
| **NIH-TCB** | | | | | | | | |
| CCC | 67.07 | 9.37 | 62.87 | 13.95 | 27 | -0.95 | .353 | -.35 |
| PV | 70.43 | 13.98 | 68.00 | 17.15 | 27 | -0.42 | .681 | -.16 |
| ORR | 68.86 | 10.81 | 63.73 | 15.01 | 27 | -1.05 | .304 | -.39 |
| PSM | 78.57 | 18.95 | 82.08 | 17.27 | 25 | 0.50 | .621 | .19 |
| PCPS | 57.07 | 8.12 | 73.21 | 22.23 | 26 | 2.55 | .017 | .97 |
| **VAS** | | | | | | | | |
| Language | 54.87 | 28.09 | 41.60 | 25.66 | 28 | -1.35 | .188 | -.49 |
| Anxiety/Irritability | 42.33 | 27.64 | 39.93 | 27.82 | 28 | -0.24 | .814 | -.09 |
| Daily Functioning | 49.93 | 29.41 | 36.13 | 24.49 | 28 | -1.39 | .173 | -.51 |
| **ABC** | | | | | | | | |
| Hyperactivity* | 3.33 | 3.02 | 6.47 | 5.89 | 20.87 | 1.83 | .081 | .67 |
| Inappropriate Speech* | 2.67 | 1.99 | 5.07 | 3.79 | 21.17 | 2.17 | .041 | .79 |
| Irritability | 5.53 | 8.12 | 7.87 | 7.43 | 28 | 0.82 | .418 | -.30 |
| Lethargy/Withdrawal | 4.13 | 5.03 | 3.33 | 4.55 | 28 | -0.46 | .651 | -.17 |
| Social Avoidance | 2.60 | 2.75 | 3.27 | 3.08 | 28 | 0.63 | .537 | .23 |
| Stereotypy | 1.80 | 3.29 | 5.07 | 5.48 | 28 | 1.98 | .058 | .72 |
| **ADAMs** | | | | | | | | |
| Depressed Mood | 1.67 | 3.11 | 2.27 | 3.37 | 28 | 0.51 | .616 | .19 |
| General Anxiety | 3.40 | 3.74 | 4.27 | 3.39 | 28 | 0.67 | .511 | .24 |
| Manic/Hyperactive Behavior | 3.07 | 2.40 | 4.73 | 3.79 | 28 | 1.44 | .161 | .53 |
| Obsessive/ Compulsive Behavior | 0.93 | 1.44 | 2.60 | 2.26 | 28 | 2.41 | .023 | .88 |
| Social Avoidance | 4.67 | 4.10 | 6.60 | 4.97 | 28 | 1.16 | .255 | .42 |

*Note.* Independent samples t-tests. Variables denoted with an asterisk (*) indicate tests that violated Levene's test for equality of variances and reported values reflect Welch's t-test statistics. All others are pooled t-test statistics.
